# What enables the safe prescription and monitoring of morphine for chronic breathlessness? Insights from a Normalisation Process Theory-informed implementation survey and interviews with clinicians

**DOI:** 10.1101/2025.06.12.25329502

**Authors:** M. Pearson, A. Mohamed, S. Bajwah, M.T Fallon, K. Date, B. Williams, D.C Currow, M.J Johnson

## Abstract

**Background:** Shortness of breath affects almost 10% of the general population, in particular people with lung or heart disease receiving palliative care. Reducing breathlessness by implementing effective pharmacological and non-pharmacological treatment is vital for relieving this physical and mental suffering in chronic illness. This mixed-methods implementation study (conducted as part of the Morphine and BrEathLessness trial (MABEL) of the clinical effectiveness and safety of regular low-dose oral modified release morphine) investigates how the experiences, attitudes and practices of clinicians affect the implementation of morphine for chronic breathlessness.

**Methods:** Mixed-methods survey and interview study with clinicians at participating trial sites, informed by Normalisation Process Theory:

1. Learning Needs Assessment survey to provide early indication of clinicians’ experiences and knowledge
2. Normalisation Measurement instrument (NoMAD) survey at two time points to measure extent to which morphine prescribing fitted with current practice
3. Semi-structured interviews with clinicians to explore perspectives about safe morphine use.

**Results:** Participants: Learning Needs Assessment (n=75, Doctors 43%, Nurses 51%, Other/not stated 6%), NoMAD 1 (n=64; Doctors 45%, Nurses 50%, Other 5%), NoMAD 2 (n=27; Doctors 41%, Nurses 52%, Other 9%), interviews (n=8, Doctors n=5, Nurses n=3). Learning Needs Assessment: two-thirds agreed they had learning needs about use of morphine for breathlessness. NoMAD surveys: 92% viewed morphine for breathlessness as a legitimate part of their role, but only 36% thought sufficient training about its use was provided. 81% stated they could integrate the use of morphine for breathlessness into their existing work, but only 39% had confidence in colleagues to do so. Interviews supported NoMAD findings and provided a richer understanding of how communication and co-ordination across settings and specialisms underpinned implementation.

**Conclusions:** Clinicians accept that morphine prescription for breathlessness management is a legitimate part of their role and are keen to improve their practice, but lack of training, resources, and confidence in the skills of others are barriers to implementation. Consistent communication about, and with, patients across settings and specialisms can enable the delivery of a safe, effective approach that enables patients to knowledgably take part in shared decision-making about the use of morphine for breathlessness.

**Trial registration:** ISRCTN87329095 (Registered 25/02/2019), EudraCT 2019-002479-33

## BACKGROUND

Shortness of breath as a part of everyday life is one of the most common forms of distress experienced by almost 10% of the general population,^1^ increasing in prevalence with age.^2^ Chronic breathlessness^3^ affects many people with lung or heart disease who are receiving palliative care.^4^ It is associated with worsening physical and mental quality of life,^5^ and challenges with all aspects of daily life^6^ including workforce participation^7^ and increased use of health services.^7^ Reducing breathlessness by implementing effective pharmacological and non-pharmacological treatment is vital for relieving this physical and mental suffering in chronic illness. The mainstay of chronic breathlessness management is addressing reversible underlying causes and then non-pharmacological interventions.^8^ The evidence base for any pharmacological intervention is strongest for morphine although the evidence is conflicting and mostly for short-term use.^9^ Knowledge about longer-term use is limited to a few studies.^10, 11^

People’s adherence to any medication is affected by treatment-related factors (including harms, dose, mode of delivery, dosing frequency), clinician factors (understanding of drug mechanism, patient-prescriber relationship), and patient factors (self-efficacy, beliefs).^12, 13, 14, 15, 16^ Despite evidence that the appropriate use of morphine is safe, it is strongly associated with clinicians’ and patients’ fears of respiratory depression, dependence and other harms.^17^

The Morphine and BrEathLessness trial (MABEL) aims to address evidence gaps around the clinical effectiveness, safety, and longer-term effects of regular low-dose oral modified release morphine for patient-reported *worst breathlessness* in people living with and needing palliation for chronic breathlessness due to advanced disease compared with placebo (Trial registration: ISRCTN87329095, EudraCT 2019-002479-33). Here, we report the findings of the trial’s linked mixed-methods implementation study, investigating how the experiences, attitudes, and practices of clinicians at participating trial sites affect the implementation of morphine for chronic breathlessness, and identifying the components of a process to support implementation of optimal prescribing and monitoring of morphine for chronic breathlessness.

## METHODS

We conducted a mixed-methods study with clinicians, informed by Normalisation Process Theory,^18, 19, 20^ comprising:

1. Learning Needs Assessment survey (see Additional File 1) - to provide an early indication of experiences and knowledge that impact on morphine prescribing.
2. Normalisation Measurement instrument (NoMAD) survey^21^ (adapted to the topic of morphine use, see Additional File 2) at two points - to measure the extent to which morphine prescribing fitted with current practice.
3. Semi-structured interviews - to explore perspectives about safe morphine use in relation to the four NPT constructs.

All surveys were administered through the secure Hull Health Trials Unit online data capture system using RedCap, consistent with an agreed data management plan. Methods and results are reported consistent with the Checklist for Reporting of Survey Studies (CROSS)^22^ and the Consolidated Criteria for Reporting Qualitative Research (COREQ)[39]. Ethical approval for the study was granted by the North East-Tyne and Wear South Research Ethics Committee (REC reference: 19/NE/0284).

### Learning Needs Assessment survey

Clinicians who were prescribers (or who supported prescribers) at each of the 12 participating sites (of which one closed before recruiting any trial participants) were invited by email to complete an anonymous online learning needs assessment prior to accessing the clinical training. In their survey responses, participants stated their learning needs on a 7-point Likert scale ranging from ‘strongly disagree’ to ‘strongly agree’ in relation to identifying patients, dose regimen, how to elicit and address patient, carer, and/or other clinician concerns, and how to manage side effects.

Completion was taken as implied consent. Participants were sent a Continuing Professional Development (CPD) certificate following completion of the subsequent 20-minute online narrated training video (delivered by a palliative care Consultant with expertise in respiratory disease, covering the current evidence base, clinical indications, dosing, management of side effects, and addressing concerns) and invited to an optional MABEL clinical training team question-answer webinar to address any remaining concerns or questions. Data were analysed descriptively (numbers and proportions) and findings fed back to the MABEL trial team to inform trial training processes.

### NoMAD surveys

On completion of the intervention training, participants were invited to complete an anonymous online modified-NoMAD survey. Prior to use, the modified survey was piloted with a clinician independent of the trial to ensure relevance, acceptability, and comprehensibility. Participants indicated their perceptions on a Likert scale regarding the extent to which the use of morphine for chronic breathlessness fitted with current practice in relation to each component of NPT - coherence with existing practices, and clinicians’ cognitive participation, collective action, and reflexive monitoring.

Completion was taken as implied consent. Participants who completed the NoMAD survey at the first timepoint and who consented to being re-contacted were invited to repeat the NoMAD survey at four to six months after their intervention training to explore changes over time. A maximum of three automated emails were sent to clinicians who had consented but not responded. Likert scale data was analysed descriptively at each timepoint (raw count (number, %) for categorical outcomes) using SPSS v.25 and Microsoft Excel, including a descriptive comparison of changes in clinicians’ perspectives pre and post-intervention. Consistent with other NPT studies and our planned descriptive analysis, a sample size calculation was not made.

### Semi-structured interviews

After completing a NoMAD survey, participants who had consented to being re-contacted about taking part in a 30-60 minute online interview (using Microsoft Teams meeting) were followed-up. Invited clinicians received a participant information sheet (study aims, what participation would involve, how anonymity would be maintained, data management, and participants’ rights), the option to discuss further with a researcher, and (for those agreeing to an interview) a consent form. Interviews were arranged for a time convenient to the participant. Each participant’s signed consent form was obtained by email prior to interview in accordance with Good Clinical Practice.

Interviews were conducted by one clinical researcher qualified in Medicine (AM) and commenced with a set of structured questions to check participants’ understanding and to collect information about the characteristics of participants and their clinical settings. The majority of each interview explored participants’ perspectives about safe morphine use through open questions in relation to the four NPT constructs. These questions, the development of which drew on the team members’ breathlessness expertise and the published literature, were used flexibly (for example, re-ordering or re-phrasing to maximise conversational flow and potential insights) as appropriate for each interview. Probes were used to highlight aspects that participants had not covered. Interviews were recorded and transcribed, read and re-read with initial reflective notes made about emerging cohesive or opposing themes. Interview transcripts were coded line-by-line using the four core constructs of NPT (coherence, cognitive participation, collective action, reflexive monitoring) and the relevant sub-component of each construct (coherence - differentiation, communal specification, individual specification, internalisation; cognitive participation - initiation, enrolment, legitimation, activation; collective action - interactional workability, relational integration, skill set workability, contextual integration; reflexive monitoring - systemisation, communal appraisal, individual appraisal, reconfiguration)^23^ using a Framework Approach^24^ in NVIVO. The coded transcripts informed the exploration of commonalities and differences between participants, moving from description towards explanation where the richness of data allowed. Initial interviews were jointly analysed by two researchers (AM and MP) with subsequent interviews analysed by AM in consultation with MP. This approach enabled the interviewing researcher (AM) to be further sensitised to arising implementation issues which were subsequently explored in later interviews, for example in relation to collective action by clinicians beyond the immediate clinical team.

## RESULTS

### Participants

The characteristics of participants in the Learning Needs Assessment, NoMAD surveys and interviews are shown in Table 1. Seventy-five clinicians (doctors 43%, nurses 51%, other/not stated 6%; 75% female) completed the Learning Needs Assessment, the majority of whom (91%) were hospital-based. Of the 89 clinicians from 12 participating sites who were invited to complete a NoMAD survey, 64 completed the initial survey (NoMAD 1) and 27 (from nine participating sites) the follow-up survey (NoMAD 2). The distribution of clinicians’ characteristics in the surveys was similar, respectively in NoMAD 1 and 2: doctors 45%, 41%; nurses 50%, 52%; other 5%, 7%; hospital-based 95%, 89%; female 75%, 81%. Of the eight interview participants, five were doctors and three were nurses. Interview participants mostly held more senior positions (four were Respiratory Consultant Physicians, two were Specialist Nurses).

**Table 1.** Characteristics of study participants, n (% (rounded))

|  | Doctor | Nurse<br>(prescriber) | Nurse (non-prescriber) | Other | Not stated | Age | Gender | Setting/ Specialty | Total |
| --- | --- | --- | --- | --- | --- | --- | --- | --- | --- |
| Learning<br>Needs<br>Survey | 32 (43%) | 9 (12%) | 29 (39%) | 4 (5%) | 1 (1%) | 21-30: 7 (9%)<br>31-40: 22 (29%)<br>41-50: 26 (35%)<br>>50: 18 (24%)<br>Not stated: 2 (3%) | Female: 56 (75%)<br>Male: 18 (24%)<br>Not stated: 1 (1%) | Hospital: 68 (91%)<br>Community: 5 (7%)<br>Hospice: 1 (1%)<br>Not stated: 1 (1%) | 75 |
| NoMAD 1 | 29 (45%) | 6 (9%) | 26 (41%) | 3 (5%) | - | 21-30: 8 (13%)<br>31-40: 18 (28%)<br>41-50: 20 (31%)<br>>50: 17 (27%)<br>Not stated: 1 (2%) | Female: 48 (75%)<br>Male: 16 (25%) | Hospital: 61 (95%)<br>Community: 2 (3%)<br>Hospice: 1 (2%) | 64 |
| NoMAD 2 | 11 (41%) | 4 (15%) | 10 (37%) | 2 (7%) | - | 21-30: 2 (7%)<br>31-40: 10 (37%)<br>41-50: 6 (22%)<br>>50: 9 (33%) | Female: 22 (81%)<br>Male: 5 (19%) | Hospital: 24 (89%)<br>Community: 2 (7%)<br>Hospice: 1 (4%) | 27 |
| Interviews | 5 | 1 | 2 | 0 | - | 21-30: 1 (13%)<br>31-40: 2 (25%)<br>41-50: 2 (25%)<br>>50: 3 (38%) | Female 5 (63%)<br>Male 3 (38%) | Respiratory -<br>Consultant: 4<br>Medicine Trainee: 1<br>Nurse Specialist: 1<br>Research Nurse: 1<br>Palliative Care Nurse: 1 | 8 |

#### Learning needs assessment survey

The results of the learning needs assessment survey are shown in Table 2. Approximately two-thirds of participants agreed that they had learning needs about the use of morphine for breathlessness in relation to identifying the person who may benefit (64%), the evidence-based dose regimen (76%), how to elicit concerns (59%), and management of side-effects (61%). A proportion of participants disagreed that they had learning needs about these areas (24%, 13%, 24%, and 24%, respectively).

**Table 2.** Learning Needs Assessment survey results (n, % (rounded))

|  | Agree | Neutral | Disagree |
| --- | --- | --- | --- |
| Regarding the use of morphine for breathlessness, I have learning needs about: |  |  |  |
| Identifying the person who may benefit | 48 (64%) | 9 (12%) | 18 (24%) |
| The evidence-based dose regimen | 57 (76%) | 8 (11%) | 10 (13%) |
| How to elicit and address patient/ carer/ other clinician concerns | 44 (59%) | 13 (17%) | 18 (24%) |
| Side-effects and how to manage them | 46 (61%) | 11 (15%) | 18 (24%) |

#### NoMAD 1 survey (immediately after intervention training)

The results of the NoMAD 1 and 2 surveys are shown in Table 3. Approximately two-thirds of participants stated that the use of morphine for breathlessness felt familiar (70%) and normal part of their clinical practice (63%), with 72% stating that they thought it could become a normal part of their clinical practice.

**Table 3.** NoMAD 1 and 2 survey results, n (% (rounded, denominator excludes respondents stating question not relevant to their role))

| Survey questions | Familiar |  | Somewhat familiar |  | Unfamiliar |  |
| --- | --- | --- | --- | --- | --- | --- |
|  | NoMAD 1 | NoMAD 2 | NoMAD 1 | NoMAD 2 | NoMAD 1 | NoMAD 2 |
| With regard to morphine for breathlessness, how <u>familiar</u> does it feel? | 45 (70%) | 22 (81%) | 12 (19%) | 4 (15%) | 7 (11%) | 1 (4%) |
| Do you feel morphine for breathlessness is <u>currently</u> a normal part of your work? | 40 (63%) | 18 (67%) | 13 (20%) | 8 (30%) | 11 (17%) | 1 (4%) |
| Do you feel morphine for breathlessness <u>will become</u> a normal part of your work? | 46 (72%) | 22 (81%) | 14 (22%) | 2 (7%) | 1 (2%) | 3 (11%) |
|  | Agree |  | Neutral |  | Disagree |  |
|  | NoMAD 1 | NoMAD 2 | NoMAD 1 | NoMAD 2 | NoMAD 1 | NoMAD 2 |
| **Coherence** |  |  |  |  |  |  |
| Staff in this organisation have a shared understanding of the purpose of morphine for breathlessness | 30 (51%) | 16 (59%) | 21 (36%) | 5 (19%) | 8 (14%) | 6 (22%) |
| I understand how morphine for breathlessness affects the nature of my own work | 52 (88%) | 24 (89%) | 7 (12%) | 1 (4%) | 0 (0%) | 2 (7%) |
| I can see the potential value of morphine for breathlessness for my own work | 60 (100%) | 27 (100%) | 0 (0%) | 0 (0%) | 0 (0%) | 0 (0%) |
| **Cognitive participation** |  |  |  |  |  |  |
| There are key people who drive appropriate use of morphine for breathlessness and get others involved | 51 (88%) | 20 (74%) | 5 (9%) | 4 (15%) | 2 (3%) | 3 (11%) |
| I believe that matters relating to morphine for breathlessness are a legitimate part of my role | 54 (92%) | 23 (85%) | 3 (5%) | 3 (11%) | 2 (3%) | 1 (4%) |
| I will continue to support appropriate use of morphine for breathlessness | 60 (98%) | 26 (100%) | 1 (2%) | 0 (0%) | 0 (0%) | 0 (0%) |
| **Collective action** |  |  |  |  |  |  |
| I can easily integrate morphine for breathlessness into my existing work | 47 (81%) | 24 (96%) | 9 (16%) | 1 (4%) | 2 (3%) | 0 (0%) |
| I have confidence in other people's ability to use morphine for breathlessness | 24 (39%) | 10 (37%) | 25 (41%) | 10 (37%) | 12 (20%) | 7 (26%) |
| Sufficient training is provided to enable staff to implement morphine for breathlessness | 21 (36%) | 5 (19%) | 16 (27%) | 10 (38%) | 22 (37%) | 11 (42%) |
| Sufficient resources are available to support morphine for breathlessness | 23 (39%) | 10 (38%) | 18 (31%) | 5 (19%) | 18 (31%) | 11 (42%) |
| Management adequately supports morphine for breathlessness | 25 (43%) | 15 (60%) | 29 (50%) | 8 (32%) | 4 (7%) | 2 (8%) |
| **Reflexive monitoring** |  |  |  |  |  |  |
| Staff agree that morphine for breathlessness is worthwhile | 42 (71%) | 17 (63%) | 17 (29%) | 9 (33%) | 0 (0%) | 1 (4%) |
| Feedback about morphine for breathlessness can be used to improve it in the future | 59 (97%) | 27 (100%) | 2 (3%) | 0 (0%) | 0 (0%) | 0 (0%) |
| I can modify how I work with morphine for breathlessness | 49 (86%) | 22 (88%) | 6 (11%) | 2 (8%) | 2 (4%) | 1 (4%) |

In relation to ***coherence***, all participants stated that they could see the potential clinical value of morphine for breathlessness, with most (88%) stating that they understood how its use would affect the nature of their own work. However, only half (51%) stated that they believed that other staff in their organisation had a shared understanding of the purpose of morphine for breathlessness.

With regard to ***cognitive participation***, most participants agreed that there were key people in their organisation who could drive appropriate use of morphine for breathlessness (88%), that matters relating to morphine for breathlessness were a legitimate part of their role (92%), and they would continue to support appropriate use of morphine for breathlessness (98%).

Regarding ***collective action***, most participants stated that they could integrate the use of morphine for breathlessness into their existing work (81% agreed, although 16% were neutral), but confidence in others was much lower with only 39% stating that they agreed that colleagues had the ability to make appropriate use of morphine for breathlessness (41% were neutral on this matter). Approximately one-third (36%) of participants agreed that sufficient training was provided to enable staff to make appropriate use of morphine for breathlessness, with 27% neutral and 37% disagreeing about the adequacy of training provision. Participants’ views about the sufficiency of resources available to support appropriate use of morphine for breathlessness (agree 39%, neutral 31%, disagree 31%) largely mirrored views about training provision. Participants’ views on management support were split, with 43% agreeing that this was adequate and 50% neutral.

In relation to ***reflexive monitoring***, most participants (97%) agreed that they could use feedback about the use of morphine for breathlessness to improve patient care. Further, they could modify how they worked in order to make appropriate use of morphine for breathlessness (86%).

#### NoMAD 2 survey (four months after intervention training)

The results of the NoMAD 1 and 2 surveys are shown in Table 3. A higher proportion of participants in the NoMAD 2 survey stated that the use of morphine for breathlessness felt familiar (81% vs. 70%), a normal part of their clinical practice (67% vs. 63%), and something that they thought could become a normal part of their clinical practice (81% vs. 72%).

NoMAD 2 results in relation to ***coherence*** were largely unchanged from NoMAD 1, with 100% (unchanged) of participants stated that they could see the potential clinical value of morphine for breathlessness, the majority (89% vs. 88%) stating that they understood how its use would affect the nature of their own work, and 59% (vs. 51%) stating that they believed that other staff in their organisation had a shared understanding of the purpose of morphine for breathlessness.

With regard to ***cognitive participation***, there was a slight fall in the proportion of participants who agreed that there were key people in their organisation who could drive appropriate use of morphine for breathlessness (74% vs. 88%) and that matters relating to morphine for breathlessness were a legitimate part of their role (85% vs. 92%). There was a small increase in the proportion who stated that they would continue to support appropriate use of morphine for breathlessness (100% vs. 98%).

Regarding ***collective action***, a higher proportion of participants in the NoMAD 2 survey stated that they could integrate the use of morphine for breathlessness into their existing work (96% vs. 81%), but confidence in others’ ability to make appropriate use of morphine for breathlessness fell slightly (37% vs. 39%). The proportion of NoMAD 2 participants who agreed that sufficient training was provided to enable staff to make appropriate use of morphine for breathlessness also fell (19% vs. 36%), with agreement on the sufficiency of resource availability to support appropriate use of morphine for breathlessness falling marginally (38% vs. 39%). Participants’ views on management support improved, with 60% (vs. 43%) agreeing that this was adequate.

In relation to ***reflexive monitoring***, participants’ views were largely unchanged, with the majority of participants agreeing that they could use feedback about the use of morphine for breathlessness to improve patient care (100% vs. 97%), that they could modify how they worked in order to make appropriate use of morphine for breathlessness (88% vs. 86%), and that appropriate use of morphine for breathlessness was worthwhile (63% vs. 71%).

#### Interviews

Interview findings are presented using the four core Normalisation Process Theory core constructs: coherence, cognitive participation, collective action, and reflexive monitoring.

### Coherence

Morphine indications and prescription depended on the situation; the use of morphine for chronic breathlessness was perceived to be different from its use as an analgesic or for chronic refractory cough. Participants prescribed morphine for pain according to the World Health Organisation (WHO) analgesic ladder (starting with non-opioid analgesics for mild pain, mild opioids for mild to moderate pain, and strong opioids for moderate to severe pain). However, this stepped approach was not used where morphine was prescribed for chronic breathlessness, but incremental strategies to *management* were used, whereby morphine was considered after other issues (e.g. hypoxaemia, anxiety) had been addressed. Morphine was considered for breathlessness due to any respiratory or cardiovascular diseases, particularly for those with advanced disease on maximal disease-related therapies.

> *“I prescribe morphine for both pain and breathlessness. I do use morphine for pain in terms of the WHO analgesic ladder. My largest experience would be in COPD because I did that COPD clinic once a week for three years.” (Respiratory consultant, 11-15 yrs of experience)*
>
> *“We tend to use morphine for chronic breathlessness. When patients have progressed further in their disease, they’re normally already on oxygen and maybe on anxiolytics as well. It just helps them to manage that sensation of breathlessness.”(Respiratory consultant, 6-10 yrs of experience)*
>
> *“I am aware that you obviously can use it with any end-stage respiratory or cardiovascular diseases.” (Registered nurse (non-prescriber), 6-10 yrs of experience)*

The mainstay of breathlessness management was seen as non-pharmacological, including pulmonary rehabilitation, following an incremental approach.

> *“I use it as an adjunct to non-medication therapies such as pulmonary rehab, breathlessness management, and fan therapy. I always use those first before. If those interventions haven’t been completely beneficial on the patient, still symptomatic, then I suggest morphine.” (Respiratory consultant, 6-10 yrs of experience)*

Practice varied between clinicians. Although most only considered morphine in the context of advanced, maximally treated disease, some felt that morphine improves lung function, reduces breathlessness, and prevents exacerbations, and could be useful in earlier-stage disease.

> *“We are thinking about giving these treatments even earlier because sometimes the patients are very breathless because you’re trying to treat them aggressively. If there is fibrosis, for example, with antibiotics, whichever becomes the only modality of treatment".(Respiratory consultant, 6-10 yrs of experience)*

There was a recognition that morphine for chronic breathlessness may help some people with chronic breathlessness, but that others stop taking it because of side-effects or lack of benefit. Some clinicians’ experience of prescribing morphine made them cautious and uncomfortable in its use because of its side-effect profile and uncertainty about any benefit, whereas others, who had seen a better harm-benefit balance in their patients were more likely to consider it. Clinicians found it difficult to predict if the person would gain benefit from the morphine or not. Worries about side-effects and addiction by both clinicians and patients (and families) were recognised. Education, reassurance, and an explanation of the reasons for prescribing morphine were essential when initiating morphine. The usual public understanding of morphine as a ‘painkiller’ in particular needed to be addressed.

> *“Some education and explanation as to why you’re prescribing morphine are required. They perceive it, of course, as a painkiller, but also recognise its addictive properties, and so it does require some education and reassurance around using it for chronic breathlessness.” (Registered nurse (prescriber) 6-10 yrs of experience)*

In a hospital setting, morphine, if prescribed, is generally for use only whilst in hospital, usually in immediate release formulations (morphine liquid, small doses) to be used ‘as required’. Clinicians usually start at low doses of morphine, with the option of increasing according to benefit and toleration. Formal measurement scales or walk tests to assess the response to morphine are not usually used. On discharge back to the community, some hospital clinicians advise the general practitioner (GP, family doctor) to prescribe a long-acting morphine based on the daily dose used in hospital, and continue morphine prescription for a therapeutic trial.

> *“We just will advise the GPs to make the prescription or if our consultants actually prescribe it during the clinic visit, then they will ask the GP to do onward repeat prescriptions.” (Associate Palliative Care Nurse, 1-2 yrs)*

If the GP and patients consider there is no benefit after three months, they are advised to stop it. Although the majority viewed morphine prescription for chronic breathlessness as common in palliative and respiratory services, consensus was not complete, with some respiratory clinicians noting that their (respiratory specialist) colleagues considered morphine to be within the purview of the palliative care team rather than respiratory physicians.

### Cognitive Participation

As described above, morphine is commonly recommended in secondary care, with GPs responsible for ongoing prescription. However, this could create problems in obtaining continuing access to morphine on hospital discharge, and options for hospital clinicians to prescribe the first month were mentioned.

> *“Having the prescription from the GP affects the logistics in terms of timely manner.” (Registered nurse (non-prescriber) 6-10 yrs of experience)*

The need to engage GPs when prescribing morphine is essential. Participants commented that historically GPs were reluctant to use morphine because they didn’t have experience of using it for chronic breathlessness. Attention to educating individual patients at the time of prescription was considered key for maintaining adherence longer-term, e.g., explaining the reason for prescription and advising the patient to get early advice about side-effects rather than stopping the medication without seeking help.

> *“Educating the patients that morphine is being used for breathlessness management, as morphine can help their condition. It is not necessarily giving it as a terminal kind of disease or illness.”* (Respiratory physician consultant, >15 yrs of experience)

Legitimisation, as part of cognitive participation, ensures that clinicians and patients believe that it is right for them to be involved and make a valid contribution (as part of their role). Therefore, education for all (clinicians, patients, family) was considered important to equip them to ensure appropriate and safe use of morphine, including co-prescribing laxatives to counter the common side-effect of constipation. It was important that all involved understood why and how morphine might help, and how a therapeutic trial should be conducted. Training sessions at conferences and other sources, including expert colleagues, were valued.

> *“There are fantastic educational sessions… at local society conferences and meetings about the importance of morphine in managing breathlessness. We all prescribe and utilize it” (Respiratory medicine trainee, 3-5 yrs of experience)*
>
> *“My specialist nurses do a lot of breathlessness, management, and education advice. I’m very lucky in the interstitial lung disease specialty that I have a pharmacist and specialist nurses. So, I’d often refer my patient to them to get counselled properly with regards to side effects, but there are no special processes in place.” (Respiratory consultant, 11-15 yrs of experience)*
>
> *“We do joint clinics and safety clinics with the palliative care consultants. That is helpful in terms of thinking about breathlessness and supporting the patients” (Respiratory consultant, 11-15 yrs of experience)*

### Collective Action

Whilst education was important, implementation strategies to facilitate putting knowledge into practice (e.g., use of care plan protocols for morphine prescription) was mentioned by some interviewees in place; a process to ensure patients are aware of the intervention, its potential benefits and side-effects. The collaboration necessary between hospital, primary and palliative care clinicians, and between doctor, nurse and allied health professionals is seen above. In the centre, the importance of joint decision making with the patient and families themselves is seen. The involvement of the extended team is commented on to ensure streamlined care:

> *“When patients are prescribed morphine for breathlessness, they are assessed by the palliative and psychology multidisciplinary team and any issues are addressed and sorted.”(Respiratory medicine trainee 3-5 yrs of experience)*
>
> *“I have a few nurse specialists that work with me, and we are comfortable to use morphine in breathlessness” Registered Nurse (non-prescriber) (Respiratory research nurse, 6-10 years)*

Involving patients in shared decision making is an essential step to maintain the intervention. Given the reported reluctance of some earlier career stage doctors and GPs regarding the use of morphine for chronic breathlessness, access to an expert clinician on the team is important. Building up confidence among the team and between the team and patients helps to implement the intervention well.

> *“We are managing well as my colleagues feel confident to care for patients with chronic breathlessness who are being managed using morphine” (Respiratory consultant, 11-15 yrs of experience)*

The division of labour to build up a set of practices to apply to the use of morphine in breathlessness is important, and requires a suitable skill mix amongst the team. This division of work appears to be led by the level of familiarity with the intervention and particular role in initiation, and ongoing prescribing. For example, interviewees found involvement of the palliative care team at initiation was a helpful step in decision making, with the GPs having more of a role in the ongoing prescribing of morphine.

> *“We’ve always made sure the GPs are very much involved in the ongoing prescribing of morphine. So, they take that on.” (Respiratory physician, 6-10 yrs of experience)*

Successful collective action was also affected by resources, policies, and procedures. Effective communication among the Multi-Disciplinary Team (MDT) helps to agree allocated resources to sustain the intervention. A closer working relationship, supported, where available, by care plans between hospital clinicians, GPs and the palliative care team enhances confidence across the ‘collective’. A lack of prescribing protocols or policies regarding morphine for breathlessness could cause confusion due to different practices.

> *“Different consultants are using different methods. Therefore, a local policy is needed.” (Respiratory consultant, 6-10 yrs of experience)*

Also, electronic prescription was identified as providing ways of flagging up potential problems and potential drug interactions, helping to improve the effectiveness and safety of the intervention. Pharmacists were highlighted as important members of the clinical team, but who may be out of the loop regarding education.

> *“The pharmacists might be less aware of the role of low dose morphine and chronic breathlessness because they’re not respiratory breathlessness specialists, they’re pharmacists. They’re drug experts.” (Respiratory consultant, 6-10 yrs of experience)*

However, having specific procedures and processes wasn’t considered important by all, with some clinicians describing morphine as just another medication on the list, with no special considerations needed.

### Reflexive Monitoring

It is clear that clinicians continually appraise the ways that using morphine for breathlessness affects their practice. The way information was collected information to determine the effectiveness of using morphine in chronic breathlessness varied. The most common approach was clinical review by a member of the clinical team.

> *“The patients might be on a 3 monthly review with a specialist nurse who would do a medication review and check if the patients still need all the medications and if they’re having any problems with it.” (Respiratory consultant, 6-10 yrs of experience)*

In addition, regular MDT meetings (usually in hospital care) were considered very important to address any concerns or problems. The community teams and nurses who assess patients on home visits were valuable, and able to report any side effects. Any benefits and problems were recorded on an electronic clinical record, although hospital and community systems may not be visible to each other. In general, clinical opinion was used with only a minority using specific questionnaires (e.g., Medical Research Council breathlessness scale) or tests (e.g., 6-minute walk test).

Appraisal of the intervention can be communal or individual appraisal. Communal appraisal shows how clinicians work together to evaluate the net benefit of using morphine in breathlessness, such as in MDT or wider oversight meetings. If there is any problem or challenge to using morphine, the clinician can bring that to this meeting.

> *“We have morbidity and mortality meetings where adverse events and things are often discussed.” (Registered Nurse (non-prescriber), 6-10 yrs of experience)*
>
> *“We have a multidisciplinary meeting once a week where we can discuss any issues related to the intervention.” (Respiratory consultant, 6-10 yrs of experience)*

Day-to-day discussions among and between clinicians, including specialists, in hospital, community and hospice settings also help in addressing the challenges that the intervention may face.

The individual appraisal indicates how a clinicians work individually to appraise the effect of the intervention and feed this into MDT discussions when reviewing patients’ progress:

> *“The community providers, the specialist nursing community, matrons, telehealth service, pulmonary rehab, other specialists, and virtual nurses all attend that meeting. We have representatives of primary care as well. So, people will sometimes come to that meeting with a question about what morphine would be appropriate for this patient, and we’ll often give advice. People can start showing more and speak about their response and whether they’re on the right dose or the right regimen and we provide advice that way.” (Respiratory consultant, 6-10 yrs of experience)*

Therefore, these meetings are important for individuals to express themselves individually to evaluate the effect of morphine on chronic breathlessness. Regular communication with the patients and taking their feedback is central to assessing the effectiveness of the intervention. Also, personal clinical supervision and regular meetings with consultants allow clinicians to reflect individually on the intervention.

As part the ongoing assessment and evaluation of the intervention, practice may be modified and procedures revised to improve its safety and effectiveness. Ongoing education is therefore needed to update clinicians. Online access appears to allow more clinicians to access training updates. Palliative care consultants were considered well-placed to provide training about how to assess and manage breathlessness, the rationale for, and when to consider morphine, its side-effects and management.

Managers need to provide time for training, and clinicians involved in setting the training agenda. Consistent with the emphasis on education, an implementation strategy based on an evidence-base ensures applying the current approach to manage breathlessness in an ongoing manner.

As well as the resources to access training, it was recognised that time resources were limited; pressure on the health service was seen as a risk to good prescribing. Interviewees felt longer appointments would allow more robust follow up with patients.

> *“We don’t have that resource in the clinic such as the logistics of having enough time to get the patient back for review in a clinic or a telephone conversation. We just don’t have that logistical capacity. So, I think that’s what is meant by the managers giving you enough time.” (Respiratory consultant, 6-10 yrs of experience)*
>
> *“We’ve got kind of six months waiting time to get an appointment at the moment. So, you just don’t have the ability to bring someone back in two or three weeks to assess the response.” (Associate Palliative Care Nurse, 1-2 yrs of experience)*

#### Synthesis of survey and interview findings

The interviews findings support the results of the NoMAD surveys, demonstrating the ways in which clinicians:

i. are open to prescribing morphine as a legitimate part of their role
ii. committed to understanding how to make appropriate use of morphine for breathlessness
iii. appreciate of the importance of careful monitoring and management of side-effects
iv. are willing to learn from authoritative sources.

The interviews also provide a richer understanding of the collective nature of making appropriate use of morphine for breathlessness and of sustaining this over time. For example, the interviews show how clinicians appreciate the importance of addressing patients’ and carers’ misconceptions about the use of morphine, of GPs’ understanding about the monitoring and maintenance of morphine in the community, and of co-ordinated communication and working processes across specialist (including palliative care) and generalist services.

## Discussion

This study describes the perspectives and attitudes of clinicians toward using morphine for chronic breathlessness. We found that hospital-based clinicians are open to learning about how to make appropriate use of morphine for breathlessness and believed this was a legitimate part of their role that they could integrate into their clinical practice. They were also open to receiving ongoing feedback about the impact of their clinical practice in relation to using morphine for chronic breathlessness. All elements of the MABEL intervention training package were welcomed and viewed as important, but only around one-third (NoMAD 1) of clinicians thought there was sufficient access to training (falling to less than a fifth in NoMAD 2), had confidence in the skills of colleagues in this context, or thought there were sufficient resources to support its safe prescription. Interview findings provided a richer understanding of how making appropriate use of morphine for breathlessness is a collective endeavour in which patients’ and carers’ misconceptions about the use of morphine also need to be addressed.

Our findings emphasising the need for education and training is consistent with other studies showing how improvement in clinicians’ knowledge and skills developed their confidence in prescribing morphine.^17, 25^ Feeling unsupported and unprepared, along with their own concerns^17^ (many of which appear to be unfounded^26^) about serious morphine-related side-effects, means that clinicians struggle to prescribe morphine and thereby fail to reach an ethical equipoise balance between the harms and benefits of morphine prescription. Supportive relationships among different clinical teams who are involved in morphine prescriptions can be achieved by enhancing trust, communication, information sharing, and understanding of clinical differences among clinicians.^17, 27^ Further, involving patients in the decision-making process is essential for safe prescriptions.^28^

We identify the importance of sufficient management support for implementing a safe prescribing and monitoring process. Although all elements of the MABEL intervention training package were welcomed and viewed as important by a wide range of clinicians involved with prescribing morphine for chronic breathlessness, there were implementation challenges due to a lack of resources.

### Strengths and limitations

We recruited a range of clinicians from participating sites to take part in the surveys and interviews, although interview participants were mostly more senior despite our efforts to engage a wider range of clinicians to participate in an interview. Our mixed-methods approach allowed both a broad and more in-depth exploration of our research questions.

The response rate for the NoMAD 2 survey was lower than for NoMAD 1, and we do not know the characteristics of the non-respondents. As this mainly affected hospital clinicians, the routine of early career doctors rotating through different departments and hospitals every 4-6 months may explain much of this fall-off. We conducted a descriptive analysis as the sample size was insufficient to support an inferential analysis. Recruiting patients and carers from the MABEL study sites for the interviews was challenging and their voices are missing - their views on the communication between hospital- and community-based clinicians and its impact on their care would have been most informative.

Finally, although numbers were much lower in the second survey, views remained stable (in particular, the positive attitude to morphine use for breathlessness). It would be interesting to survey current attitudes to morphine use in the light of a subsequent publication in a high impact journal of a trial of morphine versus placebo for chronic breathlessness due to COPD (the BEAMS trial^29^). This trial showed no evidence of benefit for breathlessness or average daily steps, which may change clinicians’ views. Although an exploratory sub-study of BEAMS gave a signal of benefit regarding levels of physical activity (inferring improved exercise endurance),^30^ this was published in a different journal and had less wide appreciation.

#### Implications for clinical practice and policy

Sufficient time and resources are required to provide opportunities for shared decision making with patients, including clear communication about i) the rationale for morphine prescription; ii) monitoring its effects; and iii) the management of side-effects. Clear and constructive interdisciplinary and multidisciplinary communication across settings and specialisms underpins the consistency of breathlessness management, and subsequently patients’ understanding. The cross-setting consistency of a safe, effective approach can be facilitated through standardised care plans and protocols.

## Conclusion

Clinicians recognise learning needs about the safe prescription and management of morphine for chronic breathlessness in practice. The experiences, knowledge, and attitudes of clinicians regarding prescribing morphine have a significant impact on implementation in clinical practice. Clinicians accept that morphine prescription for breathlessness management is a legitimate part of their role and are keen to improve their practice, but lack of training, resources, and confidence in the skills of others are barriers to implementation. Consistent communication about and with patients across settings and specialisms can enable the delivery of a safe, effective approach that enables patients to knowledgably take part in shared decision-making about the use of morphine for breathlessness.

## List of abbreviations

COPD: Chronic Obstructive Pulmonary Disease
CPD: Continuing Professional Development
GP: General Practitioner (Family Doctor)
MABEL: Morphine and BrEathLessness trial
MDT: Multi-Disciplinary Team
NoMAD: Normalisation MeAsure Development questionnaire
NPT: Normalisation Process Theory
WHO: World Health Organisation

## DECLARATIONS

### Ethics approval and consent to participate

Ethical approval for the study was granted by the North East-Tyne and Wear South Research Ethics Committee (REC reference: 19/NE/0284). Completion of surveys was taken as implied consent. Interviewees provided written informed consent to participate.

### Consent for publication

Not applicable

### Availability of data and materials

Data supporting this work are available on reasonable request. All requests will be reviewed by relevant stakeholders, based on the principles of a controlled access approach. Requests to access data should be made to Hull Health Trials Unit in the first instance.

### Authors’ contributions

MJJ, MTF, MP, SB, and DCC acquired the funding for this study. MP, SB, DCC, MJJ, and MTF conceptualised the study. MP and MJJ developed the study methodology. KD, BW, and MP provided study materials, AM collected the data, and AM, KD, BW, and MP curated the data. AM and MP analysed the data. The original draft of the manuscript was written by AM, MP, and MJJ, with review and editing by SB, MTF, KD, BW, and DCC. All authors read and approved the final manuscript.

## Acknowledgments

Our thanks to all the clinicians who participated in the training, surveys and interviews, and to all members of the MABEL Research Collaborative (Additional file 3).

## Authors information

Not applicable

