## Additional file 1 for "What enables the safe prescription and monitoring of morphine for chronic breathlessness? Insights from a Normalisation Process Theory-informed implementation survey and interviews with clinicians"

### Clinical Training

#### Learning Needs Assessment

**Thank you for attending the online clinical training for the MABEL trial.**

**This session will cover:**

1. The current evidence base regarding the:

- Importance of chronic breathlessness as a therapeutic target in its own right
- Use of oral morphine for chronic breathlessness and the rationale for the MABEL trial and its dose regimen (low dose oral modified release morphine - starting at 5mg twice daily)
- Safety profile of morphine when used for chronic breathlessness

2. Myths and fears about morphine

3. Side-effects and how to manage them

However, to make sure we cover the issues that are important for you, please take a few minutes to complete the initial learning needs survey so we can make sure we cover these in a question and answer webinar.

The group will be asked to volunteer issues which you think are important to cover.

#### Initial Learning Needs

**Please state your level of agreement with the following statements. Regarding the use of morphine for breathlessness, select the answer which most agrees with your view.**

**I have learning needs about....**

Identifying the person who may benefit

- ◯ Strongly Agree
- ◯ Agree
- ◯ Somewhat Agree
- ◯ Neither Agree nor Disagree
- ◯ Somewhat Disagree
- ◯ Disagree
- ◯ Strongly Disagree

The evidence based dose regimen

- ◯ Strongly Agree
- ◯ Agree
- ◯ Somewhat Agree
- ◯ Neither Agree nor Disagree
- ◯ Somewhat Disagree
- ◯ Disagree
- ◯ Strongly Disagree

How to elicit and address patient/carer/other clinician concerns

- ◯ Strongly Agree
- ◯ Agree
- ◯ Somewhat Agree
- ◯ Neither Agree nor Disagree
- ◯ Somewhat Disagree
- ◯ Disagree
- ◯ Strongly Disagree

Side-effects and how to manage them

- Strongly Agree
- Agree
- Somewhat Agree
- Neither Agree nor Disagree
- Somewhat Disagree
- Disagree
- Strongly Disagree

Please use the comment box to note any other areas you wish to be covered in a question and answer webinar: ____________________

#### Prescribing and Monitoring Process

**IMPORTANT INFORMATION**

As discussed we are also developing a prescribing and monitoring process for widespread use by clinicians, should the trial prove that morphine does help chronic breathlessness.

If you are happy for us to use your information to inform that process, please complete the anonymous details about you below.

There is no obligation to provide this additional part of the learning needs survey to the research team, if you decide not to participate there will not be any negative consequences; it will not affect your ability to support the MABEL study or your participation in the clinical training. You are free to decline to answer any particular question that you don't want to answer for any reason, just click next to move to the next part of the clinical training.

The responses you provide on this survey are anonymous. Although you have provided your email address to the team this will be stored separately and not linked with the information you provide here. Any information that you provide will be treated confidentially. You will not be identified individually in any project reports or publications. Your anonymous data will be held on a secure password protected computer at the Hull York Medical School. Individual responses will not be presented apart from illustrative, but anonymised, quotes from any free text. All data will be destroyed once the reports are complete.

**Indicate the most appropriate option...**

What is your age?

- ◯ 21 - 30 Years
- ◯ 31 - 40 Years
- ◯ 41 - 50 Years
- ◯ Older than 50 Years

What is your sex?

- ◯ Male
- ◯ Female
- ◯ Don't wish to say

What is your clinical profession?

- ◯ Nurse (non-prescriber)
- ◯ Nurse (prescriber)
- ◯ Doctor
- ◯ Pharmacist
- ◯ Other

Please specify 'other profession': ______________________

Jan 11, 2022 5:06:02 PM

What is your clinical setting?
*(Tick all that apply)*

- ☐ Hospital
- ☐ Community
- ☐ Hospice
- ☐ Care Home

#### Clinical Training Video

The following video consists of narrated training slides of around 20 minutes covering the evidence base for morphine in chronic breathlessness, including rationale for dose and dose schedule in the MABEL trial and practical tips on how to manage common side-effects.

Please turn on your sound.
You may expand the video size.

Please click **'Mark Training as Complete'** when you have watched the video. You will be transferred to the NOMAD survey.
