## Additional file 2 for "What enables the safe prescription and monitoring of morphine for chronic breathlessness? Insights from a Normalisation Process Theory-informed implementation survey and interviews with clinicians"

### Appendix V (NoMAD survey):



#### NOMAD Survey

##### Survey Instructions

**This survey is designed to help get a better understanding of how to apply and integrate new technologies and complex interventions in health care.**

**This survey asks questions about the implementation of oral morphine for chronic breathlessness. We understand that people involved with oral morphine prescription and monitoring for chronic breathlessness have different roles, and that people may have more than one role.**

**From the statements below please choose an option that best describes *your main role* in relation to oral morphine for chronic breathlessness.**

- ○ I am involved in identifying patients who may benefit and recommending its use
- ○ I am involved in prescribing
- ○ I am involved in monitoring for benefit or adverse effects

For this survey, please answer all the statements from the perspective of this role. Depending on your role or responsibilities, some statements may be more relevant than others.

#### Survey Instructions.

The survey is in 3 parts:

Part A asks some brief questions about yourself and your role.

Part B includes three general questions about oral morphine for chronic breathlessness (morphine for breathlessness).

Part C contains a set of more detailed questions about oral morphine for chronic breathlessness (morphine for breathlessness).

For each statement in Part C, there is the option to agree or disagree with what is being asked (OPTION A). However, if you feel that the statement is not relevant to you, there are also options to tell us why (OPTION B).

#### Part A

##### About Yourself

What is your age?

- ◯ 21 - 30 years
- ◯ 31 - 40 years
- ◯ 41 - 50 years
- ◯ Older than 50 years

What is your sex?

- ◯ Male
- ◯ Female
- ◯ Other
- ◯ Don't wish to say

What is your clinical profession?

- ◯ Nurse (non-prescriber)
- ◯ Nurse (prescriber)
- ◯ Doctor
- ◯ Pharmacist
- ◯ Other

What is your clinical setting (tick all that apply)?

- ☐ Hospital
- ☐ Community
- ☐ Hospice
- ☐ Care Homes
- ☐ Other

How many years have you worked in your current role?

- ◯ Less than one year
- ◯ 1-2 years
- ◯ 3-5 years
- ◯ 6-10 years
- ◯ 11-15 years
- ◯ More than 15 years

#### Part B

**This section asks...**

**General questions about oral morphine for chronic breathlessness (morphine for breathlessness)**

**Please indicate the most appropriate option**

#### Part B 1 of 3

With regard to morphine for breathlessness, how familiar does it feel?

- 0 - Still feels very new
- 1
- 2
- 3
- 4
- 5
- 6
- 7
- 8
- 9
- 10 - Feels completely familiar

#### Part B 2 of 3

Do you feel morphine for breathlessness is currently a normal part of your work?

- 0 - Not at all
- 1
- 2
- 3
- 4
- 5 - Somewhat
- 6
- 7
- 8
- 9
- 10 - Completely

**Part B 3 of 3**

Do you feel morphine for breathlessness will become a normal part of your work?

- 0 - Not at all
- 1
- 2
- 3
- 4
- 5 - Somewhat
- 6
- 7
- 8
- 9
- 10 - Completely

#### Part C

**This sections asks...**

**Detailed questions about oral morphine for chronic breathlessness (morphine for breathlessness)**

**For each statement please select an answer that best suits your experience using Option A.**

**If the statement is not relevant to you please select an answer from Option B.**

#### Part C 1 of 14

**For each statement please select an answer that best suits your experience using Option A. If the statement is not relevant to you please select an answer from Option B.**

Staff in this organisation have a shared understanding of the purpose of morphine for breathlessness

Option A:

- ◯ Strongly Agree
- ◯ Agree
- ◯ Neither Agree or Disagree
- ◯ Disagree
- ◯ Strongly Disagree

Option B:

- ◯ Not relevant to my role
- ◯ Not relevant at this stage
- ◯ Not relevant to the intervention

#### Part C 2 of 14

**For each statement please select an answer that best suits your experience using Option A. If the statement is not relevant to you please select an answer from Option B.**

I understand how morphine for breathlessness affects the nature of my own work

Option A:

- Strongly Agree
- Agree
- Neither Agree or Disagree
- Disagree
- Strongly Disagree

Option B:

- Not relevant to my role
- Not relevant at this stage
- Not relevant to the intervention

#### Part C 3 of 14

**For each statement please select an answer that best suits your experience using Option A. If the statement is not relevant to you please select an answer from Option B.**

I can see the potential value of morphine for breathlessness for my work

Option A:

- ◯ Strongly Agree
- ◯ Agree
- ◯ Neither Agree or Disagree
- ◯ Disagree
- ◯ Strongly Disagree

Option B:

- ◯ Not relevant to my role
- ◯ Not relevant at this stage
- ◯ Not relevant to the intervention

#### Part C 4 of 14

**For each statement please select an answer that best suits your experience using Option A. If the statement is not relevant to you please select an answer from Option B.**

There are key people who drive appropriate use of morphine for breathlessness forward and get others involved

Option A:

- ◯ Strongly Agree
- ◯ Agree
- ◯ Neither Agree or Disagree
- ◯ Disagree
- ◯ Strongly Disagree

Option B:

- ◯ Not relevant to my role
- ◯ Not relevant at this stage
- ◯ Not relevant to the intervention

#### Part C 5 of 14

**For each statement please select an answer that best suits your experience using Option A. If the statement is not relevant to you please select an answer from Option B.**

I believe that matters relating to morphine for breathlessness are a legitimate part of my role

Option A:

- ◯ Strongly Agree
- ◯ Agree
- ◯ Neither Agree or Disagree
- ◯ Disagree
- ◯ Strongly Disagree

Option B:

- ◯ Not relevant to my role
- ◯ Not relevant at this stage
- ◯ Not relevant to the intervention

#### Part C 6 of 14

**For each statement please select an answer that best suits your experience using Option A. If the statement is not relevant to you please select an answer from Option B.**

I will continue to support appropriate use of morphine for breathlessness

Option A:

- ◯ Strongly Agree
- ◯ Agree
- ◯ Neither Agree or Disagree
- ◯ Disagree
- ◯ Strongly Disagree

**Option B:**

- ◯ Not relevant to my role
- ◯ Not relevant at this stage
- ◯ Not relevant to the intervention

#### Part C 7 of 14

**For each statement please select an answer that best suits your experience using Option A. If the statement is not relevant to you please select an answer from Option B.**

I can easily integrate morphine for breathlessness into my existing work

**Option A:**

- ◯ Strongly Agree
- ◯ Agree
- ◯ Neither Agree or Disagree
- ◯ Disagree
- ◯ Strongly Disagree

Option B:

- ◯ Not relevant to my role
- ◯ Not relevant at this stage
- ◯ Not relevant to the intervention

#### Part C 8 of 14

**For each statement please select an answer that best suits your experience using Option A. If the statement is not relevant to you please select an answer from Option B.**

I have confidence in other people's ability to use morphine for breathlessness

Option A:

- ◯ Strongly Agree
- ◯ Agree
- ◯ Neither Agree or Disagree
- ◯ Disagree
- ◯ Strongly Disagree

Option B:

- ◯ Not relevant to my role
- ◯ Not relevant at this stage
- ◯ Not relevant to the intervention

#### Part C 9 of 14

**For each statement please select an answer that best suits your experience using Option A. If the statement is not relevant to you please select an answer from Option B.**

Sufficient training is provided to enable staff to implement morphine for breathlessness

Option A:

- ◯ Strongly Agree
- ◯ Agree
- ◯ Neither Agree or Disagree
- ◯ Disagree
- ◯ Strongly Disagree

Option B:

- ◯ Not relevant to my role
- ◯ Not relevant at this stage
- ◯ Not relevant to the intervention

#### Part C 10 of 14

**For each statement please select an answer that best suits your experience using Option A. If the statement is not relevant to you please select an answer from Option B.**

Sufficient resources are available to support morphine for breathlessness

Option A:

- ◯ Strongly Agree
- ◯ Agree
- ◯ Neither Agree or Disagree
- ◯ Disagree
- ◯ Strongly Disagree

Option B:

- ◯ Not relevant to my role
- ◯ Not relevant at this stage
- ◯ Not relevant to the intervention

#### Part C 11 of 14

**For each statement please select an answer that best suits your experience using Option A. If the statement is not relevant to you please select an answer from Option B.**

Management adequately supports morphine for breathlessness

Option A:

- ◯ Strongly Agree
- ◯ Agree
- ◯ Neither Agree or Disagree
- ◯ Disagree
- ◯ Strongly Disagree

Option B:

- ◯ Not relevant to my role
- ◯ Not relevant at this stage
- ◯ Not relevant to the intervention

#### Part C 12 of 14

**For each statement please select an answer that best suits your experience using Option A. If the statement is not relevant to you please select an answer from Option B.**

The staff agree that morphine for breathlessness is worthwhile

Option A:

- ◯ Strongly Agree
- ◯ Agree
- ◯ Neither Agree or Disagree
- ◯ Disagree
- ◯ Strongly Disagree

Option B:

- ◯ Not relevant to my role
- ◯ Not relevant at this stage
- ◯ Not relevant to the intervention

#### Part C 13 of 14

**For each statement please select an answer that best suits your experience using Option A. If the statement is not relevant to you please select an answer from Option B.**

Feedback about morphine for breathlessness can be used to improve it in the future

Option A:

- ◯ Strongly Agree
- ◯ Agree
- ◯ Neither Agree or Disagree
- ◯ Disagree
- ◯ Strongly Disagree

Option B:

- ◯ Not relevant to my role
- ◯ Not relevant at this stage
- ◯ Not relevant to the intervention

#### Part C 14 of 14

**For each statement please select an answer that best suits your experience using Option A. If the statement is not relevant to you please select an answer from Option B.**

I can modify how I work with morphine for breathlessness

Option A:

- ◯ Strongly Agree
- ◯ Agree
- ◯ Neither Agree or Disagree
- ◯ Disagree
- ◯ Strongly Disagree

Option B:

- ◯ Not relevant to my role
- ◯ Not relevant at this stage
- ◯ Not relevant to the intervention

#### Follow Up Consent

**If you would be willing to repeat this survey in about 4 months, so we can see how opinions might change over time, please answer 'Yes' to the consent question:**

I consent to repeat the survey in about 4 months

- ○ Yes
- ○ No

Are you happy for us to contact you about taking part in an interview about your experiences of morphine prescription and breathlessness?

- ○ Yes
- ○ No
