## Additional file 3 for "What enables the safe prescription and monitoring of morphine for chronic breathlessness? Insights from a Normalisation Process Theory-informed implementation survey and interviews with clinicians"

### MABEL Research Collaborative

| Key role | Title | First name | Surname | Highest degree | Organisation, country |
|---|---|---|---|---|---|
| Chief Investigator | Prof | Marie | Fallon | MD | Edinburgh Cancer Centre<br>MRC Institute of Genetics & Molecular Medicine, The University of Edinburgh, Western General Hospital, University of Edinburgh. UK |
| Chief investigator | Prof | Miriam | Johnson | MD | Wolfson Palliative Care Research Centre, Hull York Medical School, University of Hull, UK |
| Co-Investigator / Site Principal Investigator | Dr | Sabrina | Bajwah | PhD | Cicely Saunders Institute of Palliative Care, Policy & Rehabilitation, Florence Nightingale Faculty of Nursing, Midwifery & Palliative Care, King's College London, London. UK |
| Co-Investigator / Site Principal Investigator* | Dr | Nazia | Chaudhuri | MD | Faculty of Life & Health Sciences, School of Medicine, University of Ulster, Northern Ireland, UK |
| Co-Investigator | Prof | David | Currow | PhD | University Technology Sydney, Sydney, Australia |
| Co-Investigator | Prof | Judith | Cohen | PhD | Hull Health Trials Unit, Hull York Medical School, University of Hull, UK |
| Co-Investigator / Site Principal Investigator | Prof | Simon | Hart | PhD | Respiratory Medicine Clinical Trials Unit, Hull York Medical School, University of Hull, UK |
| Co-Investigator | Prof | Irene | Higginson | PhD | Cicely Saunders Institute of Palliative Care, Policy & Rehabilitation, Florence Nightingale Faculty of Nursing, Midwifery & Palliative Care, King's College London, London. UK |
| Co-Investigator** | Prof | John | Norrie | MSc | Centre for Public Health, Queen's University Belfast, Belfast, Northern Ireland, UK |
| Co-investigator | Dr | Mark | Pearson | PhD | Wolfson Palliative Care Research Centre, Hull York Medical School, University of Hull, UK |
| Trial Manager | Ms | Bronwen | Williams | MSc | Hull Health Trials Unit, Hull York Medical School, University of Hull, UK |
| Statistician | Ms | Catriona | Keerie | MSc | Edinburgh Clinical Trials Research Unit, University of Edinburgh, UK |
| Statistician | Mrs | Sharon | Tuck | MSc | Edinburgh Clinical Trials Research Unit, University of Edinburgh, UK |
| Health Economist | Dr | Peter | Hall | PhD | Edinburgh Cancer Centre, University of Edinburgh, UK |
| Health Economist | Mr | Marek | Atter | MA | Edinburgh Cancer Centre, University of Edinburgh, UK |

*Manchester site PI until 24 March 2022; **formerly Edinburgh Clinical Trials Unit, University of Edinburgh, UK

### Other MABEL Research Collaborative members

| Key role | Title | First name | Surname | Organisation |
|---|---|---|---|---|
| Site Principal Investigator | Prof | Rachael | Evans | University Hospitals of Leicester NHS Trust, NIHR Leicester Biomedical Research Centre - Respiratory, Glenfield Hospital, Leicester. |
| Site Principal Investigator | Dr | Shaney | Barrett | North Bristol NHS Trust, Trust Headquarters, Southmead Hospital, Bristol, |
| Site Principal Investigator | Dr | Hannah | Bayes | NHS Greater Glasgow & Clyde, Glasgow Royal Infirmary, Glasgow |
| Site Principal Investigator | Dr | Gareth | Stewart | NHS Lothian, Western General Hospital, Edinburgh |
| Site Principal Investigator | Dr | Gareth | Walters | Birmingham University Hospitals Birmingham NHS Foundation Trust, Birmingham |
| Site Principal Investigator* | Dr | Pilar | Rivera-Ortega | NIHR Manchester Clinical Research Facility, Wythenshawe Hospital, Manchester University NHS Foundation Trust, Manchester |
| Site Principal Investigator | Mr | Jonathon | Palmer | University Hospitals Plymouth NHS Trust, Derriford, Plymouth |
| Site Principal Investigator | Dr | Alison | Boland | Leeds Teaching Hospital NHS Trust, St James University Hospital, Leeds |
| Site Principal Investigator | Dr | Joanna | Bowden | NHS Fife, Queen Margaret Hospital, Dunfermline, Scotland |
| Patient and Public Involvement | Mrs | Annie | Jones | INVOLVE Hull, University of Hull |

***From 30 March 2022**

#### Acknowledgements

We are grateful to all the patients and caregivers who took part in this study. We also want to acknowledge the helpful advice and input from our patient and public involvement (PPI) group members.

We thank the members of the independent Data Monitoring and Ethics Committee (including Professor Brian Le, Professor Paddy Stone, Rhian Gabe); Trial Steering Committee (Professor Anthony Byrne, Dr Helen Mossop, Dr David Meads, and PPI members Konstantin Kamenev and Pam Mackay).

We thank:
• Sponsor Representatives – Mr James Illingworth, Ms Leanne Cox, Hull University Teaching Hospital's NHS Trust.
• Hull Health Trials Unit - HULL, UK: Ms Bronwen Williams, Mr Anesu Matamba, Dr Matthew Northgraves, Mr Paul Bradley, Mrs Charlotte Thompson, Mrs Sarah Sumpter, Mrs Beccy Acaster, Ms Amy Porter, Mrs Kerri Morris
• Health Economics – Dr Peter Hall, Marek Atter, University of Edinburgh
• Statisticians – Ms Catriona Keerie, Ms Sharon Tuck, University of Edinburgh, Edinburgh Clinical Trials Unit
• Other clinicians, including recruiting clinicians and research nurses in particular: University Hospitals Birmingham NHS Trust, UK: Siobhan Wilkinson, Louise Wood, Babita Rajkumari; Castle Hill Hospital, UK: Rachel Flockton, Caroline Wright, Rachel Thompson; Leeds Teaching Hospitals NHS Trust: Dr Suzie Gillon, Jodie Glossop, Gaynor Martin, Clair Favager; Western General Hospital, NHS Lothian, Edinburgh, UK: Dr Rebecca Dickinson, Jaqueline Henderson; NHS Fife, UK: Dr Devesh Dhasmana, Julie Penman, Emma Simpson, Anne Marie De La Santos, Amanda Pratt, Angela Scullion; Glasgow Royal Infirmary, UK: Dr David Anderson, Charlene Dunne; King's Hospital, UK, UK: Paramjote Kaler, Stefania Stegner; NIHR Manchester Clinical Research Facility, Wythenshawe Hospital, Manchester, UK Janet Johnston, Sue Stockdale; University Hospitals of Leicester NHS Trust, UK: Dr Thomas Ward, Dr Lorna Latimer, Kate Porter, Emily Morgan-Selvaratnam, BRC Respiratory theme, Dept of Respiratory Sciences, University of Leicester; University Hospital NHS Trust Plymouth; Jonathon Palmer, Memory Mwadeyi; University Hospitals Bristol NHS Foundation Trust, Caroline Kilby, Nuria Novas Duarte
